# Biofilm Derived Oxylipin Mediated Autoimmune Response in Breast Implant Subjects

**DOI:** 10.1101/2020.11.18.20233510

**Authors:** Imran Khan, Robert E. Minto, Christine Kelley-Patteson, Bruce W. Van Natta, Colby R. Neumann, Lily J. Suh, Kanhaiya Singh, Mary Lester, R Jason VonDerHaar, Gayle M. Gordillo, Aladdin H. Hassanein, Chandan K. Sen, Marshall E. Kadin, Mithun Sinha

**Author notes:** Corresponding Author: Mithun Sinha, PhD, Address: 975 W Walnut St, Medical Research Library Building, Suite # 444A, Indiana University School of Medicine, Indianapolis, IN 46202.

## Abstract

Over 10 million women worldwide have breast implants for breast cancer/prophylactic reconstruction or cosmetic augmentation. In recent years, a number of patients have described a constellation of symptoms that are believed to be related to their breast implants. This constellation of symptoms has been named Breast Implant Illness (BII). The symptoms described include chronic fatigue, joint pain, muscle pain and a host of other manifestations often associated with autoimmune illnesses. In this work, we report that bacterial biofilm is associated with BII. We postulate that the pathogenesis of BII is mediated *via* a host-pathogen interaction whereby the biofilm bacteria *Staphylococcus epidermidis* interacts with breast lipids to form the oxylipin 10-HOME. The oxylipin 10-HOME was found to activate CD4^+^ T cells to Th1 subtype. An increased abundance of CD4^+^Th1 was observed in the breast tissue of BII subjects. The identification of a mechanism of immune activation associated with BII via a biofilm enabled pathway provides insight into the pathogenesis for implant-associated autoimmune symptoms.

## Introduction

Breast implants were first introduced in 1962. Nearly 60 years later, their safety has continued to be controversial in the medical field, including a period of FDA-mandated restrictions on the use of silicone gel breast implants in the U.S. in the 1990s^1, 2^. Nearly 300,000 women have breast implant surgeries every year in the United States, for reasons including cosmetic augmentation, post-mastectomy breast reconstruction (breast cancer and prophylactic mastectomy), revision of prior augmentation/ reconstruction, and gender affirmation^3^. A subset of patients with breast implants have a myriad of nonspecific systemic symptoms^4, 5^. The symptoms described include fever, myalgias, chronic fatigue, arthralgias and a host of other manifestations often associated with autoimmune illnesses^6-12^. This constellation of symptoms related to implants has been named Breast Implant Illness (BII). The number of patients who opt for breast implant explantation due to complications including breast implant illness is over 30,000 annually^3^. Thus, BII is a growing concern to both patients and surgeons alike with more than 10 million women worldwide currently having breast implants^6^. Despite the increased concern regarding BII, existing scientific literature doesn’t show a definite link between breast implants and autoimmune or connective tissue diseases. Some epidemiological studies have reported that women with silicone gel-filled implants were 8 times more likely to be diagnosed with Sjögren syndrome, an autoimmune disorder characterized by dry eyes and a dry mouth. Also, such subjects are 7 times more likely to be diagnosed with scleroderma, a group of autoimmune diseases that cause the skin and connective tissues to become hard and tighten, and nearly 6 times more likely to be diagnosed with rheumatoid arthritis^8^. Other studies have reported an association of autoimmune symptoms with breast implants^7, 9-12^. Symptoms have been documented to begin after placement of the implant and are often relieved by their explantation ^13, 14^. This led patients and physicians to suspect that the breast implants are the likely cause ^13, 14^. However, FDA mandated studies have found silicone gel breast implants to be safe^15^. Of note to this apparent contradiction is the fact that these symptoms have been reported in subjects with other implants such as orthopedic arthroplasty which is typically comprised of titanium^16-20^. This indicates that the underlying cause of these conditions may be associated with factors other than the implant material. Therefore, it is important to decipher the underlying molecular mechanism associated with BII for a better understanding in the future of all implant-related illnesses with autoimmune manifestations.

This is the first in depth study to investigate the possible role of bacterial biofilm as a factor in the pathogenesis of BII through a patient-based study with a mechanistic pathway. We have identified a biofilm-derived oxylipin in increased abundance in BII subjects. The oxylipin led to activation of CD4^+^ Th1 cells in an *in vitro* and *in vivo* mouse model suggesting a role in the establishment of an autoimmune response often observed to be associated with BII.

## Results

### Bacterial Biofilm in Implant-Associated with BII

The study included 68 patients. Forty-six patients with BII were analyzed. Subjects were diagnosed with BII using clinical parameters outlined in previous studies ^6-12^ **(Fig. 1A)**. As a part of the diagnosis, the patients were required to complete a questionnaire **(Supplementary Table 1)**. The questionnaire screened for the commonly reported symptoms associated with BII ^6-12^. Implant, associated capsules and breast tissue were collected from these subjects **(Fig. 1B-C)** as described in methods. The mean age of BII patients was 44.0 years with mean duration of implant insertion of 11.51 years. Two groups were considered as controls. Control group I (non-BII, n=14) was comprised of patients with breast implants who didn’t exhibit BII symptoms but went through explantation of the breast implant. The mean age of non-BII patients was 46.5 years. Control group II (normal tissue, n=8) was comprised of patients without an implant, whose breast tissue was removed as a clinically indicated surgical procedures e.g., (breast reduction surgery, mammoplasty, contralateral procedures for reconstruction symmetry and mastopexy). The mean age of normal subjects was 26.8 years. The demographics of the subjects have been provided in **Supplementary Table 2**.

**Figure 1.**
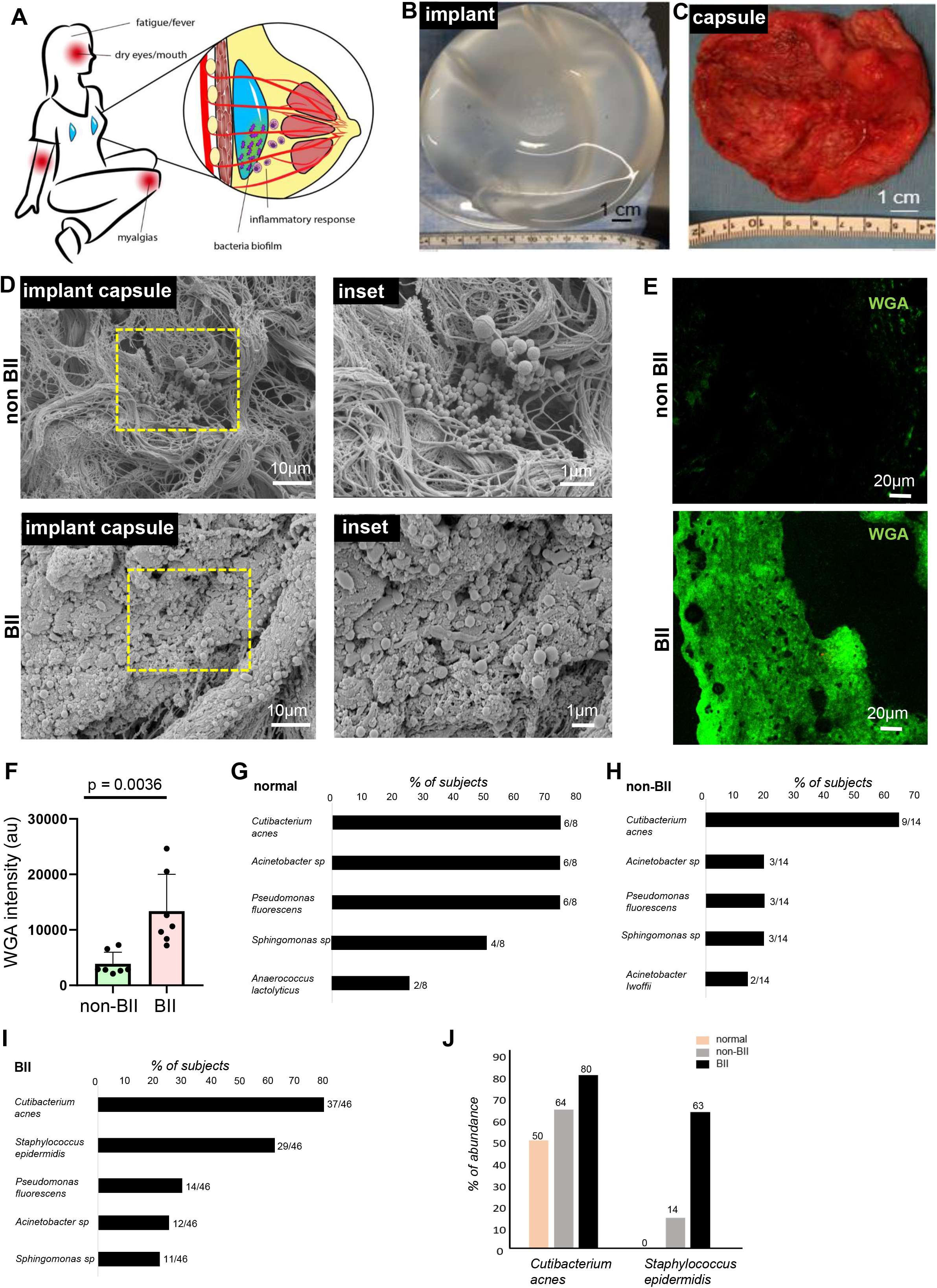
Bacterial Biofilm in Implant-Associated with BII. **(A)** Schematic presentation of the bacterial biofilm association with breast implant illness (BII) **(B)** Breast implant isolated from a subject **(C)** Capsule associated with breast implant of the subject shown in (B) **(D)** Increased abundance of bacterial biofilm from the implant-associated capsule of BII compared to non BII subjects as determined through scanning electron microscopy. Zoomed insets of region of interest (ROI) dotted yellow square shown. n=10 non-BII subjects, n=25 BII subjects. **(E-F) E**, Increased abundance of bacterial biofilm as measured through wheat germ agglutinin (WGA) assay in the capsules of BII subjects compared to the non-BII capsules. **F**, quantification of biofilm aggregates using WGA staining. Data presented as mean ± SEM, n=7 non-BII, n=7 BII **(G-I)** 16S rRNA NGS based bacterial typing from the breast tissues of **G**, normal; **H**, non-BII and **I**, BII subjects. Top five bacterial species in each group represented. Absolute values of subject samples associated with a bacterial species is provided in parenthesis n=8 (normal), n=14 (non-BII), n=46 (BII) **(J)** Increased abundance of biofilm forming *Staphylococcus epidermidis* in implant-associated breast tissues of BII subjects. Percentage of Patients with *Cutibacterium acnes* and *Staphylococcus epidermidis* provided above the individual bars. n=8 (normal), n=14 (non-BII), n=46 (BII)

Bacterial biofilm was observed in implant-associated capsules through scanning electron microscopy **(Fig. 1D, Supplementary Fig. S1A)**. Though biofilm was detected in the capsules of implants from both BII and non-BII patients, the abundance of biofilm was significantly more in the capsules of BII subjects as observed through wheat germ agglutin (WGA) assay **(Fig 1E-F)**. The microbiological culture analyses of the tissues resulted in no growth of bacterial colonies (data not shown). It has been reported that bacterial biofilms are difficult to detect through colony forming assays due to their subdued metabolism ^21, 22^. Hence, in cases of bacterial biofilm, next generation sequencing (NGS) using variable region of bacterial 16S rRNA gene are employed to type bacteria and determine their abundance^23^. Diverse types of biofilm forming bacteria were observed associated with BII, non-BII and normal tissues **(Fig. 1G-I)** through next generation sequencing (NGS) of the 16S rRNA variable region. Most of the species identified were opportunistic bacteria associated with normal skin flora capable of forming biofilms. **(Fig. 1G-I)**. Comparative analyses with the normal, non-BII and BII groups revealed an increased abundance of *Staphylococcus epidermidis* in BII **(Fig. 1J)**. The other bacteria found in increased abundance in breast tissue was *Cutibacterium acnes*. Bivariate analysis using cross-tabulation was performed between presence of biofilm and the study groups. Using the Fisher’s exact test, *Staphylococcus epidermidis* colonization was observed to be significantly higher (p<0.001) in the BII group (63.04%) compared to non-BII group (14.3%) and the normal group. Using exact logistic regression, the BII group was 9.8 times significantly more likely to have *S. epidermidis* colonization compared to the non-BII group (p=0.003). Similarly, when comparing with normal groups, the BII group was 17.4 times significantly more likely to have *S. epidermidis* (p=0.0021). *C. acnes* was not found to be associated (Fisher’s exact p-value=0.116) with the groups defined in the study. However, though not significant, the BII group (80.4%) tended to show 4 times higher odds of colonization with *C*.*acnes* (Odds ratio from exact logistic regression=3.98 [95% CI: 0.32, 26.10, p=0.1684]) compared to the normal (50.0%) group **(Fig. 1J, Supplementary Fig. S1B)**.

### Increased Abundance of Biofilm Derived 10-HOME in BII Subjects

The oxylipin (10)-hydroxy-(8*E*)-octadecenoic acid (10-HOME) is formed by the bacterial oxidation of oleic acid **(Fig. 2A)**. The oxylipin 10-HOME has been reported to inhibit flagellum-driven swimming and swarming motilities and stimulate the formation of bacterial biofilms *in vitro*^24^. The oxylipin 10-HOME was synthesized in the laboratory (commercially not available) in natural isotopic abundance (light) isotope and deuterated (heavy) isotope forms. The former was used for biological experiments, whereas the later was used as a liquid chromatography-mass spectrometry (LC-MS) standard. The synthesized 10-HOME was validated through thin layer chromatography and NMR spectroscopy **(Supplementary Fig. S2A-B)**.

**Figure 2.**
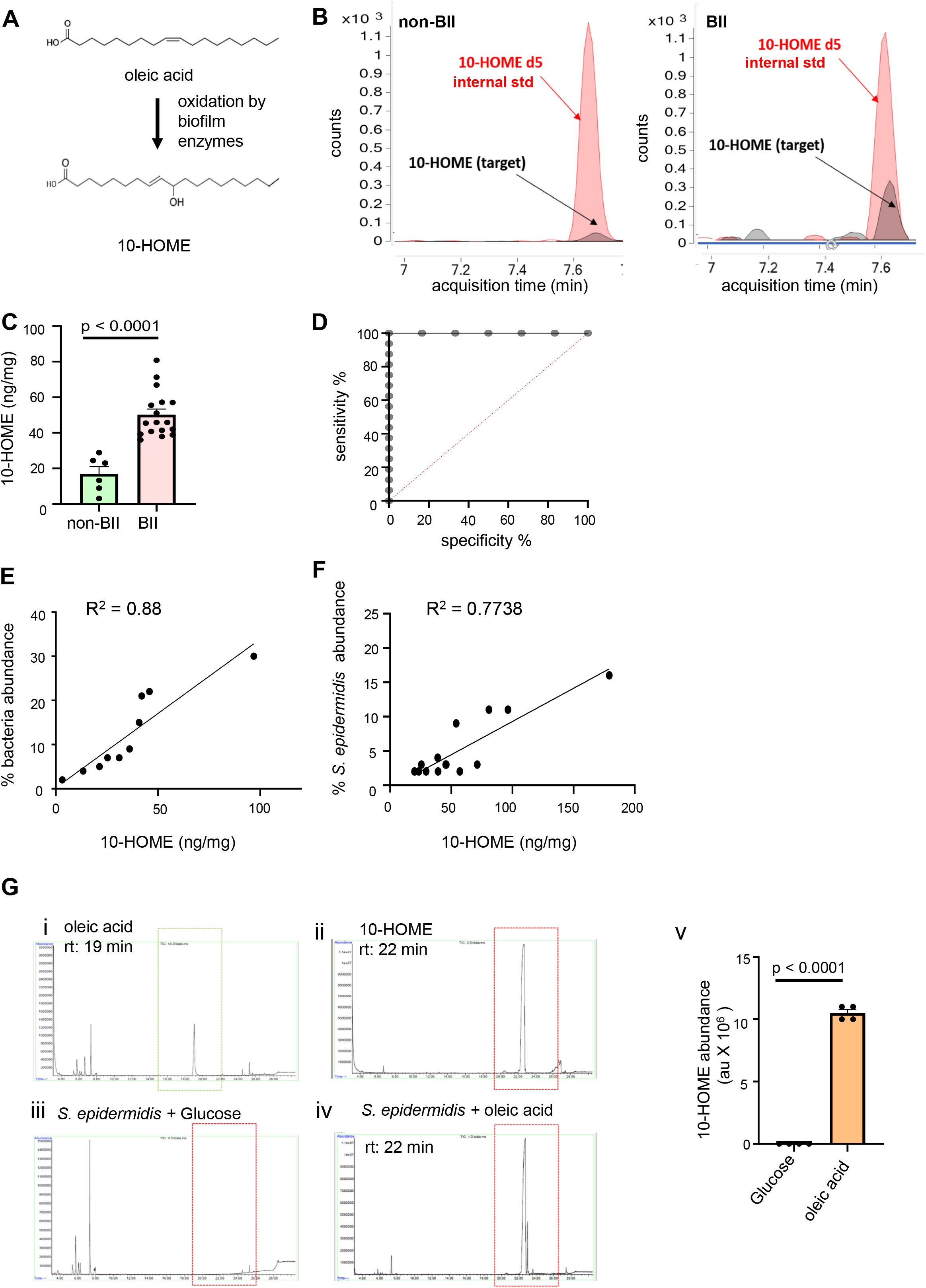
Increased Abundance of Biofilm Derived 10-HOME in BII Subjects. **(A)** Schematic of formation of 10-HOME from oleic acid **(B-D)** Increased abundance of 10-HOME in implant associated tissue of BII subjects. **B**. Chromatograms of 10-HOME from non-BII and BII using LC-MS/MS targeted analyses. **C**. Data presented as mean ± SEM, n=6 (non-BII), n=17 (BII). **D**. Receiver operating characteristic (ROC) curve analysis to determine specificity and sensitivity of 10-HOME detection. **(E)** Increased abundance of bacteria associated with 10-HOME detected from the implant associated tissue of BII subjects. **(F)** Increased abundance of *Staphylococcus epidermidis* associated with 10-HOME detected from the implant associated tissue of BII subjects. **(G)** Synthesis of 10-HOME by *S. epidermidis in vitro* upon using oleic acid as carbon source. Gas chromatography-mass spectrometry analyses for detection of 10-HOME. **i**-oleic acid standard, **ii**-10-HOME standard, **iii**-*S*.*epidermidis* with glucose as carbon source, **iv**-*S*.*epidermidis* with Oleic acid as carbon source, **v**-quantification of 10-HOME abundance. n=4

Elevated levels of 10-HOME in implant-associated samples of BII compared to non-BII samples were observed through mass spectrometry (**Fig. 2B-D**). Positive correlation was observed between bacterial abundance and concentration of 10-HOME in BII subjects ***(Fig. 2E)***. Similar correlation was observed in BII subjects with *Staphylococcus epidermidis* **(Fig. 2F)**. To determine if *S. epidermidis* was capable of synthesizing 10-HOME, it was cultured *in vitro* with oleic acid as a source of carbon. Formation of 10-HOME was detected using gas chromatography-mass spectrometry **(Fig. 2G)**. Oxylipins have been reported to cause CD4^+^ T cell activation^25^. Hence, we explored the role of CD4^+^ T cells in BII.

### Abundance of CD4^+^ Th1 Cells in Implant-Associated Tissue of BII Subjects

Breast tissue associated with the implant of BII subjects showed increased presence of CD4^+^ T-BET^+^ T cells compared to that of non-BII breast tissue **(Fig. 3A)**. T-BET transcription factor is critical in Th1 subtype determination^26^. Th1 cells are associated with an auto-immune response in multiple illnesses including rheumatoid arthritis^27^. The CD4^+^ Th1 cells associated with BII subjects also were CD36^+^ **(Fig. 3B)**. CD36 is a fatty acyl translocase^28^; its level is upregulated when uptake of fatty acid (normal or oxidized) is required. It is to be noted that BII is a systemic autoimmune manifestation. Hence, the peripheral blood of BII and non-BII subjects was analyzed for CD4^+^ T cells (Th1, Th2, Th9 and Th22). Increased abundance of Th1 cells was observed in BII subjects (N=9) as measured through flow cytometry using CD183/CXCR3 (CD4^+^ Th1 cell marker) **(Fig. 3C)**. No significant difference was observed in the abundance of other Th subtypes Th2 (CD194), Th9 (CD196) and Th22 (CD196) between BII subjects and non-BII subjects **(Supplementary Fig. S3A-B)**. The following human cell lines were used as positive control for validation of surface antigens. Mac2A for CD183, Mac2B for CD194^29^ and TLBR1 for CD196^30^ **(Supplementary Fig. S3C-E)**. These results, however, don’t definitively establish that 10-HOME led to CD4^+^Th1 cell induction or that 10-HOME can lead to CD36 upregulation. Thus, we studied the effect of 10-HOME on human primary naïve CD4^+^ T cells.

**Figure 3.**
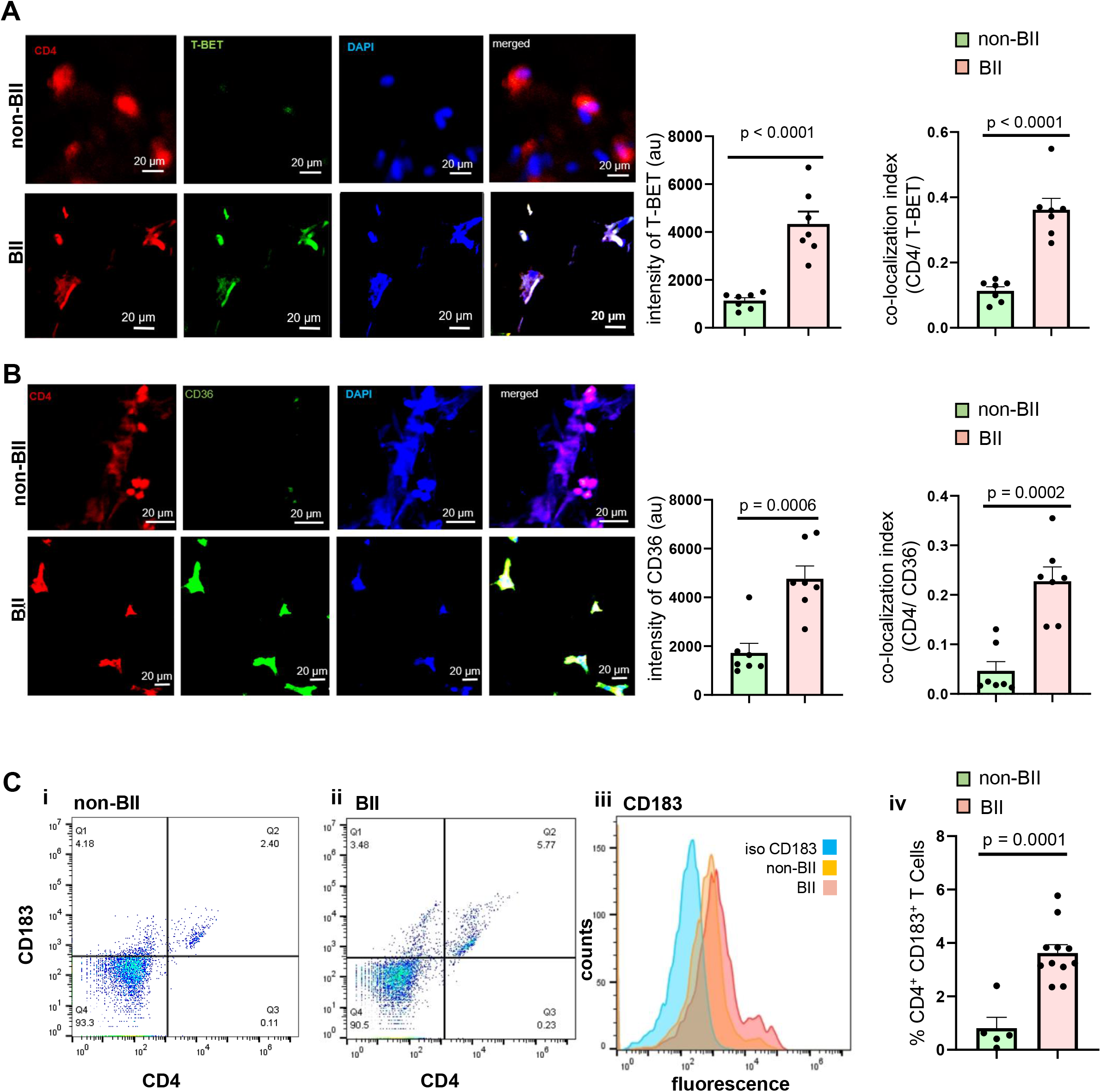
Abundance of CD4+ Th1 Cells in Implant Associated Tissue of BII Subjects. **(A)** Increased expression of T-BET (transcription factor for Th1 subtype) in breast tissue associated with BII subjects compared to non-BII breast tissue. Breast tissue immuno-stained with anti-CD4 (red) antibody, anti-T-BET (green) and DAPI (blue). Intensity of TBET for the two groups, colocalization of CD4 and T-BET depicted increased abundance of CD4^+^ TBET^+^ cells in the breast tissues of BII subjects. Data presented as mean ± SEM, (n=7). Scale bar = 20 μm **(B)** Increased expression of CD36 (fatty acyl translocase) in breast tissue associated with BII subjects compared to non-BII breast tissue. Breast tissue immuno-stained with anti-CD4 (red) antibody, anti-CD36 (green) and DAPI (blue). Intensity of CD36 for the two groups, colocalization of CD4 and CD36 depicted increased abundance of CD4^+^ CD36^+^ cells in the breast tissues of BII subjects. Data presented as mean ± SEM, (n=7). Scale bar = 20 μm **(C)** Elevated Th1 subtype in the peripheral blood of BII subjects. Flow cytometry analyses of peripheral blood of subjects stained with anti-CD4 (FITC) and anti-CD183 (PE). Representative flow plots. (i) non-BII (ii) BII (iii) histogram with isotype control for CD183 (iv) % of CD4^+^ CD183^+^ T cells. Data presented as mean ± SEM, (n=5-11).

### Oxylipin 10-HOME Polarizes Naïve CD4+ T Cells to Th1 Subtype *in vitro*

In order to study the effect of 10-HOME on T cells, naïve CD4^+^ T cells (isolated from human peripheral blood mononuclear cells) were treated with 100 µM 10-HOME for 48h. Increased CD36 expression was observed in the 10-HOME treated CD4^+^ T cells through immunocytochemistry **(Fig. 4A)**, flow cytometry **(Fig. 4B)** and quantitative real time PCR **(Fig. 4H)** indicative of the 10-HOME mediated induction of CD36. Polarization to Th1 subtype occurred in the presence of 10-HOME as observed through immunocytochemistry **(Fig. 4C)**, flow cytometry **(Fig. 4D)**, and quantitative real time PCR **(Fig. 4I)**. The CD4^+^ cells exhibited increased expression of T-BET (a transcription factor activated during the polarization of naïve T cells to Th1 subtype^26^)(**Fig. 4C, 4H, Supplementary Fig. 4A**), CD183/ CXCR3 (CD4^+^ Th1 cell marker) (**Fig. 4D**), and Th1 secreted pro-inflammatory cytokine IFN-γ through ELISA (**Fig. 4E**). The CD183^+^ Th1 cells exhibited increased abundance of CD36 marker (**Supplementary Fig. 4B**). The other subtypes of CD4^+^ T cells (Th2, Th9 and Th22) assayed didn’t exhibit any statistically significant increase in abundance post 10-HOME treatment on naïve CD4^+^ T cells. Th2 cells were assayed using surface marker CD194/CCR4 (**Supplementary Fig. 4C**), transcription factor GATA3 (**Supplementary Fig. 4D**) and anti-inflammatory cytokines IL 4 and IL10 (**Fig. 4F-G**). Th9 and Th22 cells were assayed using surface marker CD196/CCR6 (**Supplementary Fig. 4E**).

**Figure 4.**
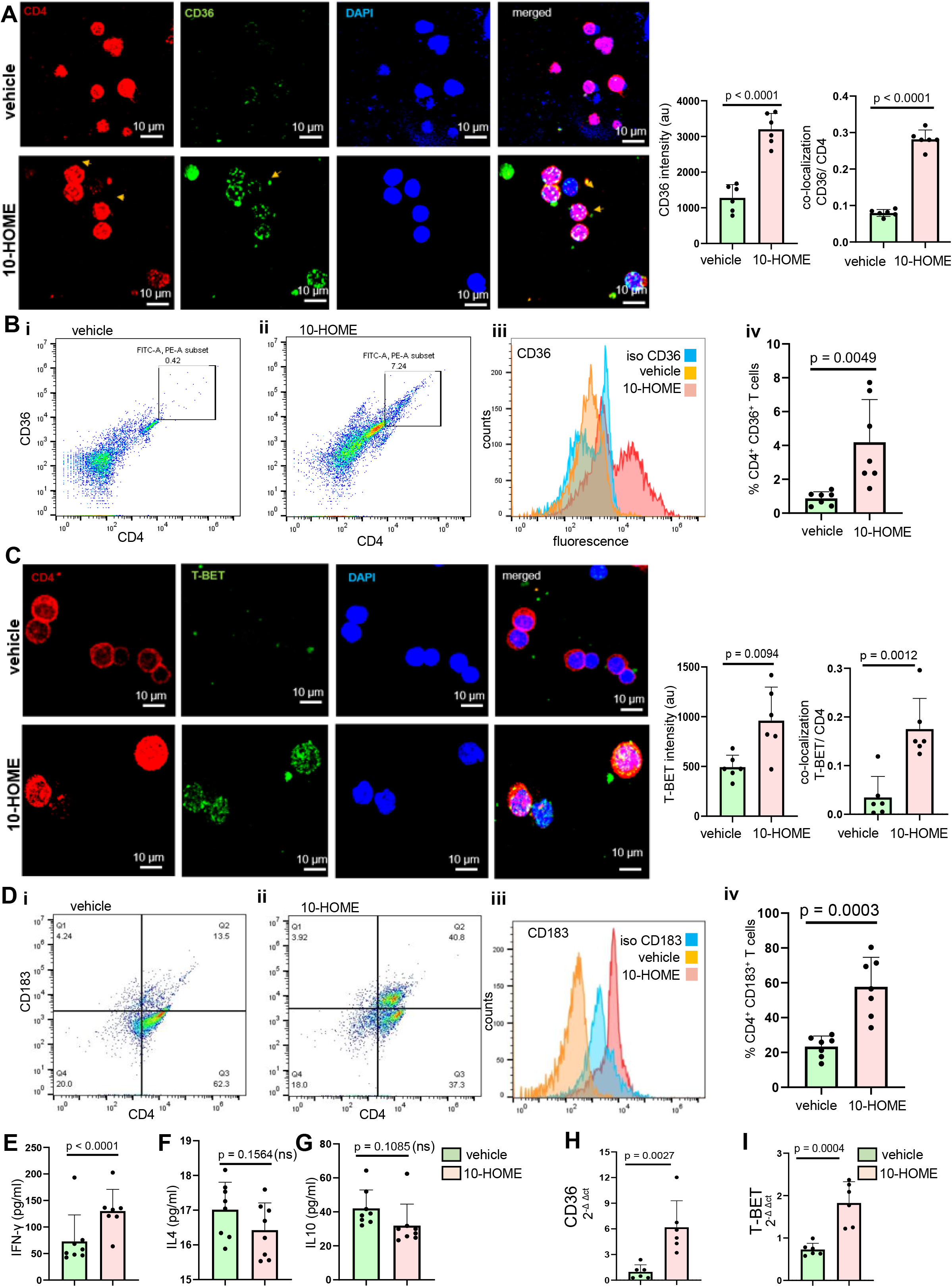
Oxylipin 10-HOME Polarizes Naïve CD4+ T Cells to Th1 Subtype *in vitro*. **(A)** Increased expression of CD36 in naïve CD4^+^ T cells treated with 100 µm of 10-HOME compared to vehicle post-48h. T cells were immuno-stained with anti-CD4 (red) antibody, anti-CD36 (green) and DAPI (blue). Intensity of CD36 quantified for the two groups. Colocalization of CD4 and TBET depicted increased abundance of CD4^+^ CD36^+^ cells in the breast tissues of BII subjects. Data presented as mean ± SD, (n=6). Scale bar = 10 μm **(B)** Elevated CD36 in the 10-HOME treated naïve T cells. Flow cytometry analyses of treated cells stained with anti-CD4 (FITC) and anti-CD36 (APC). Representative flow plots. (i) vehicle treated (ii) 10-HOME treated (iii) histogram with isotype control for CD36 (iv) % of CD4^+^ CD36^+^ T cells. Data presented as mean ± SD, (n=7). **(C)** Increased expression of T-BET in naïve CD4^+^ T cells treated with 100 µm of 10-HOME compared to vehicle post-48h. T cells were immuno-stained with anti-CD4 (red) antibody, anti-TBET (green) and DAPI (blue). Intensity of TBET quantified for the two groups. Colocalization of CD4 and TBET depicted increased abundance of CD4^+^ TBET^+^ cells in the breast tissues of BII subjects. Data presented as mean ± SD, (n=6). Scale bar = 10 μm **(D)** Elevated Th1 subtype (CD183^+^) in the 10-HOME treated naïve CD4^+^ T cells. Flow cytometry analyses of treated cells stained with anti-CD4 (FITC) and anti-CD183 (PE). Representative flow plots. (i) vehicle treated (ii) 10-HOME treated (iii) histogram with isotype control for CD183 (iv) % of CD4^+^ CD183^+^. Data presented as mean ± SD, (n=7). **(E)** Increased expression of Th1 secreted pro-inflammatory cytokine IFN-γ in the 10-HOME treated naïve CD4^+^ T cells as measured through ELISA. Data presented as mean ± SD (n=7). **(F-G)** No significant change in the cytokines IL4 and IL10 following10-HOME treatment of naïve CD4^+^ T cells as measured through ELISA, Data presented as mean ± SD, (n=8). **(H-I)** Increased expression of (H) CD36 (I) T-BET in 10-HOME treated naïve T cells as measured through quantitative real time PCR. Data presented as mean ± SD, (n=6).

### Elevated CD4^+^ Th1 in Peripheral Blood of Mice Administered with 10-HOME

To test if 10-HOME can induce Th1 cells *in vivo*, we administered 10-HOME into the abdominal mammary fat pad of mice (**Fig. 5A**) as described in the Methods. An increased abundance of CD4^+^ Th1 cells was found in the murine blood 14 days post administration of 10-HOME (**Fig. 5B**). Other subtypes of CD4^+^ T cells (Th2) didn’t exhibit any statistically significant increase in abundance **(Fig. 5C)**.

**Figure 5.**
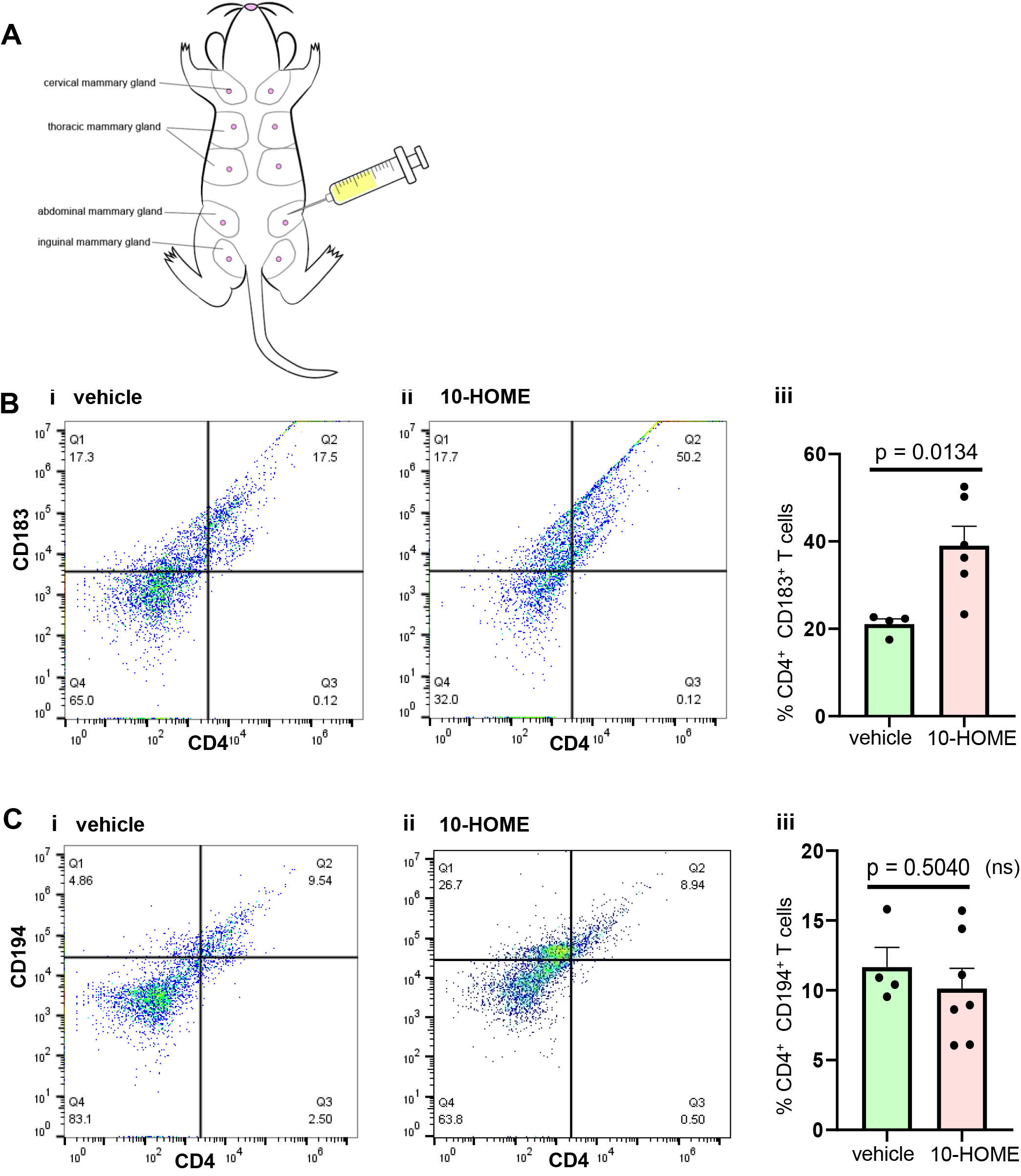
Elevated CD4^+^ Th1 in Peripheral Blood of Mice Administered with 10-HOME. **(A)** Schematic representation of injection of 10-HOME in the abdominal mammary fat pad of mice. **(B)** Elevated CD4^+^ Th1 subtype in the blood of mice injected with 10-HOME. Flow cytometry analyses of blood of mice stained with anti-CD4 (FITC) and anti-CD183 (PE). Representative flow plots. (i) Vehicle treated (ii) 10-HOME treated (iii) % of CD4^+^ CD183^+^ T cells. Data presented as mean ± SEM, (n=4-6 mice). **(C)** Unaltered CD4^+^ Th2 subtype in the blood of mice injected with 10-HOME. Flow cytometry analyses of blood of mice stained with anti-CD4 (FITC) and anti-CD194 (PE). Representative flow plots. (i) Vehicle treated (ii) 10-HOME treated (iii) % of CD4^+^ CD194^+^ T cells. Data presented as mean ± SEM, (n=4-6 mice).

## Discussion

Biofilm formation enables single-cell microbes to assume a temporary multicellular lifestyle, in which “group behavior” facilitates survival in adverse environments^31^. The transition from planktonic growth (associated with acute pathogenic infections) to biofilm occurs in response to environmental changes, and involves multiple regulatory networks thereby mediating the spatial and temporal reorganization of the bacterial cell^31^. Bacterial biofilms have been implicated to cause gastric cancer by *Helicobacter pylori*^32^, colon cancer^33, 34^, breast implant-associated anaplastic large cell lymphoma (BIA-ALCL) and chronic inflammation^35-37^. The factors that involve interplay between host and pathogen are influenced by the micro-environmental niche where the bacteria reside^22, 38, 39^. Breast implants provided a conducive surface for the adherence and growth of bacterial biofilms^40^. The breast is a modified sweat gland, with multiple external opening from the mammary ducts *via* the nipple, providing a passage for microbes (normal skin flora) to the breast tissue. This concept has been supported by a report whereby researchers have shown the presence of microbes deep inside the breast tissue^41^. Many bacteria belonging to the normal microflora of the body have been reported to form bacterial biofilms^42^. *Pseudomonas aeruginosa*^43, 44^, *Staphylococcus aureus* ^45^ and *Ralstonia picketii* ^40, 46^ are some of the common biofilm forming species associated with breast implants. The bacterial biofilms are difficult to detect since they have subdued metabolism and may not be detectable as CFUs in standard microbiological assays ^21, 22^. They can be identified through structural assays like scanning electron microscopy (SEM)^22, 47^ and genetic assays (e.g.,16S rRNA sequencing)^48^. The observation in this study of increased abundance of bacterial biofilm comprised of *Staphylococcus epidermidis* associated with implant-associated tissue of BII patients relative to controls was thus critical in understanding to the potential pathology of BII. It is to be noted that while this study was being performed, anecdotal evidence of *S*.*epidermidis* with BII was reported by Mark *et al* ^49^.

The oxidation of fatty acids is one of the main biochemical reactions in the synthesis of lipid mediators. The oxygenation of unsaturated fatty acids leads to the formation of oxylipins, although free fatty acids are not readily available as they are found as triglycerides. The action of bacterial lipases, such as dioxygenases (DOX) and lipoxygenases (LOX) on breast adipose result in the availability of free fatty acids. When oleic acid is used as a bacterial substrate, it is oxidized to form 10-HOME. The unique adipose tissue found in breast tissue is rich in oleic acid containing lipids^50, 51^. The oxylipin 10-HOME has been reported to promote bacterial biofilms *in vitro*^24^. The increased abundance of 10-HOME in our study associated with breast tissues of BII subjects thus provides evidence that breast microflora may interact with breast lipids promoting the formation of bacterial biofilms.

Oxylipins are also known to be immune-modulatory. It has been reported that 12,13 DiHOME derived from oxidation of linoleic acid led to the reduction of regulatory T cells (Tregs), impeded immune tolerance and promoted childhood atopy and asthma^25^. Elevated levels of 10-HOME formed due to bacterial biofilms could lead to immune cell activation. Reports suggest that CD4^+^ T cells are activated due to the persistent presence of a bacterial biofilm^52, 53^. CD4^+^ T cells play an important role in the pathogenesis of chronic systemic inflammatory autoimmune diseases such as multiple sclerosis, diabetes and rheumatoid arthritis^54-56^. CD4^+^ T cells can be divided into Th1, Th2, Th9, Th17 and Th22 subsets based upon the cytokines they produce. In chronic inflammatory systemic autoimmune diseases such as diabetes, multiple sclerosis and rheumatoid arthritis, Th1 cells were found to be involved^57^. It is to be noted that many of the symptoms of BII patients were similar to rheumatoid arthritis and multiple sclerosis^8^. Th1 cells trigger delayed type hypersensitivity (DTH) reactions (which are mostly mediated by macrophages) and immunoglobulin class switch towards IgG2a^55, 56^.

Oxidized lipids have been associated with pain and inflammatory conditions^58^. Pain reported as arthralgia and myalgia are common in BII. We identified increased presence of CD4^+^ Th1 cells in the breast tissue and peripheral blood of BII subjects. Findings of this study demonstrate that 10-HOME was capable of polarizing naïve CD4^+^ T cells towards Th1 subtype *in vitro*. Increased abundance of transcription factor T-BET required for Th1 fate was identified post 10-HOME treatment. The polarization to Th1 subtype was also supported by the observation of increased expression of pro-inflammatory cytokine IFN-γ secreted by Th1 cells. Systemic analyses in our study didn’t revealed polarization towards Th2, Th9 and Th22 subtypes. It is to be noted that the current work does not rule out the polarization of naïve CD4^+^ T cells to Th17 subtype upon treatment with 10-HOME. Th17 subtype has been also reported to be associated with auto-immune syndrome^59^.

To correlate the association of 10-HOME with activation of CD4^+^ T cells *in vivo*, mice were administered with 10-HOME in their mammary fat pad. An increased abundance of CD4^+^ Th1 cells in peripheral circulation in 10-HOME administered mice was observed. These observations also help to explain the increased abundance CD4^+^ Th1 in the BII subjects.

Taken together, we investigated the biofilm hypothesis of breast implant illness through a host-pathogen interaction. The breast microenvironment led to formation of biofilm derived 10-HOME from host oleic acid. The 10-HOME thus formed led to activation of CD4^+^ Th1 cells *in vitro* and *in vivo*. The study provides the first evidence of a possible link of biofilm derived 10-HOME inducing an autoimmune response in patients with BII. Additionally, in light of reports of biofilm association with metal implants such as orthopedic arthroplasty, this study provides an explanation of autoimmune responses reported using those metal implants^16-20^. The findings of this study suggest that management of biofilm can help to increase the safety and long-term use of implants. Further research needs to be conducted to elucidate if other biofilm forming bacterial species are involved in the pathogenesis of BII. Also, more investigation is warranted to decipher the pathway leading to the onset of BII post-biofilm formation and the involvement of other biofilm derived metabolites.

## Materials and Methods

### Human subjects

Subjects participating in the study were patients diagnosed with BII. Demographic characteristics of patients presented in **(Supplementary Table 2)**. All human studies were approved by The Indiana University School of Medicine Institutional Review Board. Declaration of Helsinki protocols was followed, and patients gave their written informed consent.

### Animals

All animal (mice) experiments were approved by the Indiana University School of Medicine Institutional Animal Care and Use Committee (SoM-IACUC) under protocol 19102 - Murine model of breast implant diseases. Animals were housed under a 12-h light–dark cycle with food and water *ad libitum*.

### Bacterial strains

Staphylococcus epidermidis (Winslow and Winslow) Evans (ATCC® 35984™) were grown on tryptic soy agar plate at 37°C.

### Scanning Electron Microscope Imaging

The samples were collected in glutaraldehyde fixation buffer, dehydrated with graded ethanol, and treated with hexamethyldisilazane (HMDS, Ted Pella Inc.) and left overnight for drying^22, 38^. Before scanning, samples were mounted and coated with gold. Samples were imaged with FEI™NOVA nanoSEM scanning electron microscope (FEI™, Hillsboro, OR) equipped with a field-emission gun electron source.

### Wheat-germ agglutinin (WGA) staining

Paraffin embedded capsules surrounding the implant were sectioned on the slide. Wheat Germ Agglutinin, Alexa Fluor™488 Conjugate (Invitrogen) stock solution (1mg/ml) was diluted in PBS. The sections were stained with Wheat Germ Agglutinin (dilution 1:200) for 10 mins ^60^. The slides were then mounted and imaged on a Zeiss LSM 880 microscope equipped with the AIRYscan detector.

### NGS sequencing for 16S rRNA

Microbial DNA in each sample were sequenced by MicrogenDx Inc using the Illumina MiSeq sequencer. Forward and reverse primers were used to detect and amplify the target sequence, for 16S gene in bacteria. The samples are differentiated from each other when run on the MiSeq sequencer by a “tag,” a unique identifying sequence attached to the forward and reverse primers implemented when the targeted sequence is amplified using PCR. Following PCR, purification of the pooled DNA was done by removing small fragments using both Agencourt Ampure beads and Qiagen Minelute kit. The DNA was quantified and prepared for sequencing. Finally, the DNA library was run on the MiSeq sequencer. The sequencing reads were analyzed for quality and length during the data analysis. The data analysis pipeline consisted of two major stages, the denoising and chimera detection stage and the microbial diversity analysis stage. During the denoising and chimera detection stage, denoising was performed using various techniques to remove short sequences, singleton sequences, and noisy reads. With the low-quality reads removed, chimera detection was performed to aid in the removal of chimeric sequences. The high-quality sequencing reads of the variable region of 16S rRNA were compared to curated database of MicrogenDx. The database is comprised of 18500 unique bacteria.

### Synthesis of 10-HOME

For the synthesis of 10-HOME, a convergent Horner-Wadsworth-Emmons approach was employed. Indiana University has filed a provisional patent application (Application # 63/107,626) on behalf of REM, IK and MS relating to the methods and synthesis of 10-HOME and its deuterated 17,17,18,18,18 d_5_ analog to be used as analytical standards.

### Primary T-cell isolation

Naïve CD4^+^ T cells were isolated from peripheral blood mononuclear cells (PBMC). Briefly, PBMCs were isolated by Ficoll-Paque PLUS density gradient sedimentation. Naïve CD4^+^ T cells were then enriched using immunomagnetic, column-free, negative selection (Naïve CD4 T cell isolation kit, Miltenyi Biotec)^61^. Unwanted cells (CD8, CD14, CD15, CD16, CD19, CD25, CD34, CD36, CD45RO, CD56, CD123, TCRγ/δ, HLA-DR, and CD235a (Glycophorin A)) were removed using antibody complexes recognizing non-naïve CD4 T cells and dextran-coated magnetic particles.

### Primary CD4^+^ T cell culture and 10-HOME treatment

Primary CD4^+^ T cells were cultured under standard conditions at 37°C in a humidified incubator with 5% CO _2_ in RPMI-1640 growth medium supplemented with 10% FBS, 100 IU/ml penicillin, 0.1 mg/ml streptomycin, 10 mmol/l L-glutamine supplemented with IL2 for 48h^62^. Following that, CD4^+^ T cells were treated with oxylipin 10-HOME (100 µM) or vehicle control for 48 h.

### Immunohistochemistry and immunocytochemistry

Paraffin embedded breast tissue blocks were sectioned, deparaffinized and immunostained^22, 39, 63^. Immunohistochemical staining of the sections were performed using standard procedures using the following primary antibodies: α-CD4 antibody (abcam# ab133616; dilution 1:200), α-CD36 antibody (Abcam # ab80080, clone MF3, dilution: 1:200), α-T-bet antibody (Abcam # ab91109, clone 4B10, dilution: 1:200). To enable fluorescence detection, sections were incubated with appropriate Alexa Fluor® 488 (green, Molecular probes), or Alexa Fluor® 564 (red, Molecular probes) conjugated secondary antibodies. The sections were counterstained with DAPI (Sigma). For immunocytochemistry, cells were fixed with IC fixation buffer (eBioscience), blocked with 10 percent normal goat serum (Vector Laboratories), incubated with primary and secondary antibodies and counterstained with DAPI. Mosaic images were collected using a Zeiss Axiovert 200 M, inverted fluorescence microscopy or confocal microscopy (LSM880). Image analysis was performed using Zen (Zeiss) software to quantitate fluorescence intensity (fluorescent pixels).

### Lipid extraction and 10-HOME quantification using LCMS

LC-MS/MS targeted analysis from capsule and breast adipose tissue was performed. Samples were weighed and transferred to 2 ml vials with 1.4 mm ceramic beads and 1 ml of water with 0.1% formic acid. The standard solution was prepared by aliquoting 1µl of each stock solution into a new tube drying the original solvent and solubilizing in 1 ml of 100% ethanol to obtain a final concentration of 1 ng/ml each. Samples were homogenized using Precellys24 tissue homogenizer (Bertin Technologies, Rockville, MD). The total volume of the homogenate was extracted with ethyl acetate in a 1:1 volume ratio. Samples were vortexed for 1 minute and centrifuged at 14.000 rpm for 10 minutes. The organic phase was collected and transferred to a new vial to be evaporated and stored at −80°C until analysis. The dried lipid extracts were reconstituted with 50 μl of methanol/water at 1:1 volume ratio and submitted for targeted quantification by liquid chromatography tandem MS (LC/MS/MS). The LC column used was an Acquity UPLC BEH C18 1.7µm particle size - 2.1×100 mm (Waters, Milford, MA). The binary pump flow rate was set at 0.3mL/min in an Agilent UPLC (G7120A) using water and 0.1% formic acid as mobile phase A and acetonitrile and 0.1% formic acid as mobile phase B. The LC column was pre-equilibrated with 80% A for 1 min. The binary pump was set in a linear gradient to 100% B in 8 min and held for 2.50 min. It was then returned to 80% A and re-equilibrated for 4 min. Ten μL of the reconstituted sample was delivered to the column through a multisampler (G7167B) into a QQQ6470A triple quadrupole mass spectrometer (Agilent Technologies, San Jose, CA) equipped with ESI Jet Stream ion source. In the mass spectrometer the capillary voltage was 3500 V on the negative ion mode, the gas temperature was 325°C, gas flow was set at 8l/min, the sheath gas heater at 250°C and the sheath gas flow at 7 l/min. The fragmentation voltage was 100 and the cell accelerator voltage was 4 V. The MRMs (parent-fragment) for the acquisition included were 297.5->155.4 for 10-HOME and for the internal standard it was 302.4->155.4. Concentrations in ng/mg of tissue were obtained by normalizing by the dried weight of the sample homogenized and by the concentration of the deuterated internal standard. To quantify 10-HOME, calibration curves were done with 7 serial dilutions of the stock solution starting at 10 ng as the highest concentration. Data processing was carried out by using Mass Hunter (B.06.00) using software Quant and Qual.

### Flow cytometry analyses

The fluorescence and light-scattering properties (forward scatter and side scatter) of the cells were determined by using BD Accuri C6. Signals from cells labeled with conjugated fluorophores were detected. The following antibodies were used for different flow cytometry analysis. PE anti-human CD183 (CXCR3) (clone G025H7, Biolegend # 353705, 2 μg/ml), PE anti-human CD194 (CCR4) (clone L291H4, Biolegend # 359412, 0.5 μg/ml), PE anti-human CD196 (CCR6) (clone G034E3, Biolegend # 353410, 0.5 μg/ml), FITC anti-human CD4 (clone A161A1, Biolegend # 357406, 0.5 μg/ml), APC anti-human CD194 (CCR4) (clone L291H4, Biolegend # 359407, 0.5 μg/ml), Alexa Fluor® 700 anti-human CD196 (CCR6) (clone G034E3, Biolegend # 353433, 0.5 μg/ml), PE anti-human CD36 (clone 5-271, Biolegend # 336205, 1 μg/ml), FITC anti-mouse CD4 (clone GK1.5, Biolegend # 100406, 0.5 μg/ml), APC anti-mouse CD36 (clone HM36, Biolegend # 102612, 0.5 μg/ml), PE anti-mouse CD183 CD36 (clone CXCR3-173, Biolegend # 126506, 0.5 μg/ml), PE anti-mouse CD196 (clone 29-2L17, Biolegend # 129804, 0.5 μg/ml), APC anti-mouse CD194 (clone 2G12, Biolegend # 131211, 0.5 μg/ml), PE anti-mouse FOXP3 (clone MF14, Biolegend # 126403, 0.5 μg/ml), PE anti-T-bet Antibody (clone 4B10, Biolegend # 644809, 0.5 μg/ml). For intracellular markers, TBET and GATA3 permeabilization was performed through True-nuclear transcription factor buffer set (Biolegend # 424401). Auto compensation was performed using samples stained with single flurophores. Gates were set manually. BD Diva (BD Biosciences), and FlowJo softwares were used for analyses^63^. Logarithmic scale was used to measure cell fluorescence. Appropriate IgG control fluorescence compensation was applied to avoid false positive signals.

### ELISA

Cell-free supernatants were collected and stored at −80°C. ELISAs for IFN-γ, IL4 and IL10 were performed using DuoSet kits (R&D Systems) as per manufacturer’s protocol.

### 10-HOME administration in mice

At approximately 9 to 10 weeks of age, female mice were anesthetized with isoflurane, and five injections of 10-HOME (15 mg/kg body weight) every alternate day was made with a 27 G needle to the abdominal mammary fat pad of mice. Post-two weeks of the dose the mice were euthanized. Blood, mammary fat pad and spleen were harvested for subsequent analyses.

### Quantitative RT PCR

Breast tissue was pulverized using tissue pulverizer (6770 Freezer/Mill) and total RNA was extracted using miRVana (Thermo Fisher Scientific). cDNA was made using SuperScript™III First-Strand Synthesis System (Invitrogen) or SuperScript™VILO™cDNA Synthesis Kit (Invitrogen). Quantitative or real-time PCR (Sybr Green) approach was used for mRNA quantification ^63-68^. Primer sequences used in this study provided in (**Supplementary Table 3**).

### Quantification and statistical analysis

The data analysis was performed using student’s t-test (two-tailed) to determine significant differences. Mean, standard deviation and student paired *t*-test analysis was done using in-built function in Microsoft Excel 2010. Data are presented as mean ± SEM (*in vivo*) or ± SD (*in vitro*) as reported in figure legends. Comparisons among multiple groups were tested using ANOVA in-built function in GraphPad Prism 8.4.2. p<0.05 was considered statistically significant.

## Data Availability

All relevant data are available from the corresponding author upon reasonable request.

## ACKNOWLEDGEMENTS

This work was supported by Aesthetic Surgery Education and Research Foundation (ASERF) grant to MS, Department of Defense PR200435, and American Association of Plastic Surgeons grants to AHH, the US National Institutes of Health grants NIH NR015676, U01DK119099 to GG and CKS; NIH R01GM108014, R01NS042617, R01DK125835 to CKS. The authors acknowledge Core Services for SEM at INDI Core, Indiana University Purdue University Indianapolis. We thank Metabolite Profiling Facility, Purdue University, West Lafayette. We thank Dr. Lava Timsina, Indiana University School of Medicine for his help with the statistics. We thank Dr. Allen Epstein, Keck School of Medicine, University of Southern California for his kind donation of the breast implant-associated anaplastic large cell lymphoma (BIA-ALCL), T cell line TLBR1.

## AUTHOR CONTRIBUTIONS

MS, IK, and AHH conceived and designed the work. IK, REM, CKP, BWV, CRN, LS, KS, ML, JVD participated in the data acquisition. MS, REM, IK, CKP, AHH and MEK wrote the manuscript. MS, REM, CKS, GMG, MEK, BWV reviewed the manuscript.

