## Supplementary file for "Biofilm Derived Oxylipin Mediated Autoimmune Response in Breast Implant Subjects"

A

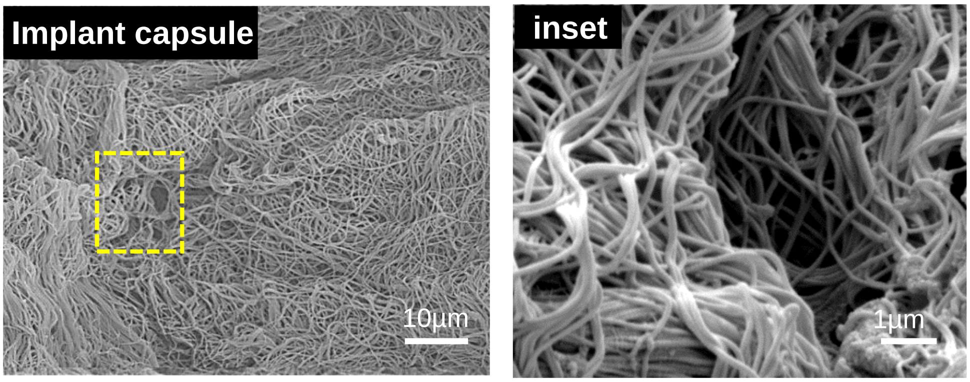

B

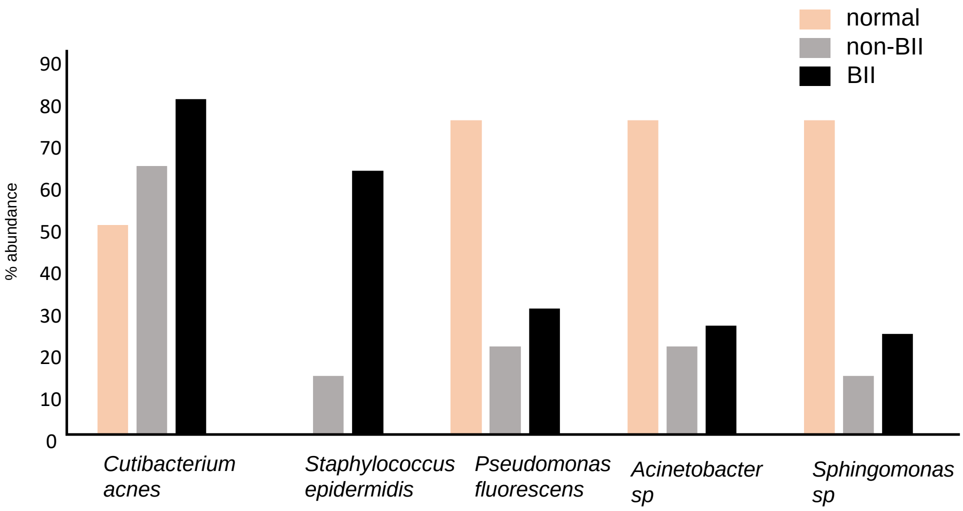

**Supplementary Figure 1**

**Supplementary Fig 1**

**(A)** Scanning electron microscopy and zoomed inset showing the collagen fibers of a implant-associated capsule from a non-BII subject. The subject didn't exhibit biofilm in the capsule.

**(B)** Abundance of NGS identified top five bacterial species from the tissues of normal, non-BII and BII subjects. n=8 (normal), n=14 (non-BII), n=46 (BII)

**A**

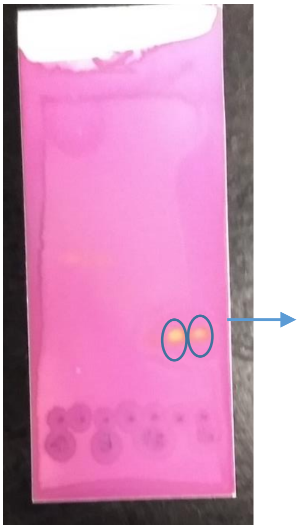

(10S)-hydroxy-(8E)-octadecenoic acid (10-HOME)

**B**

(10)-hydroxy-(8E)-octadecenoic acid ($C_{18}H_{34}O_3$)

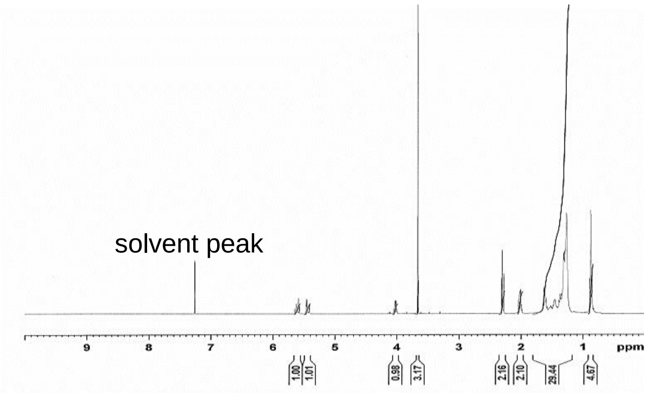

**Supplementary Figure 2**

**Supplementary Fig 2**

**(A)** 10-HOME (internal standard) synthesis validation using thin-layer chromatography (TLC)

**(B)** Validation of 10-HOME standard using proton nuclear magnetic resonance (NMR)

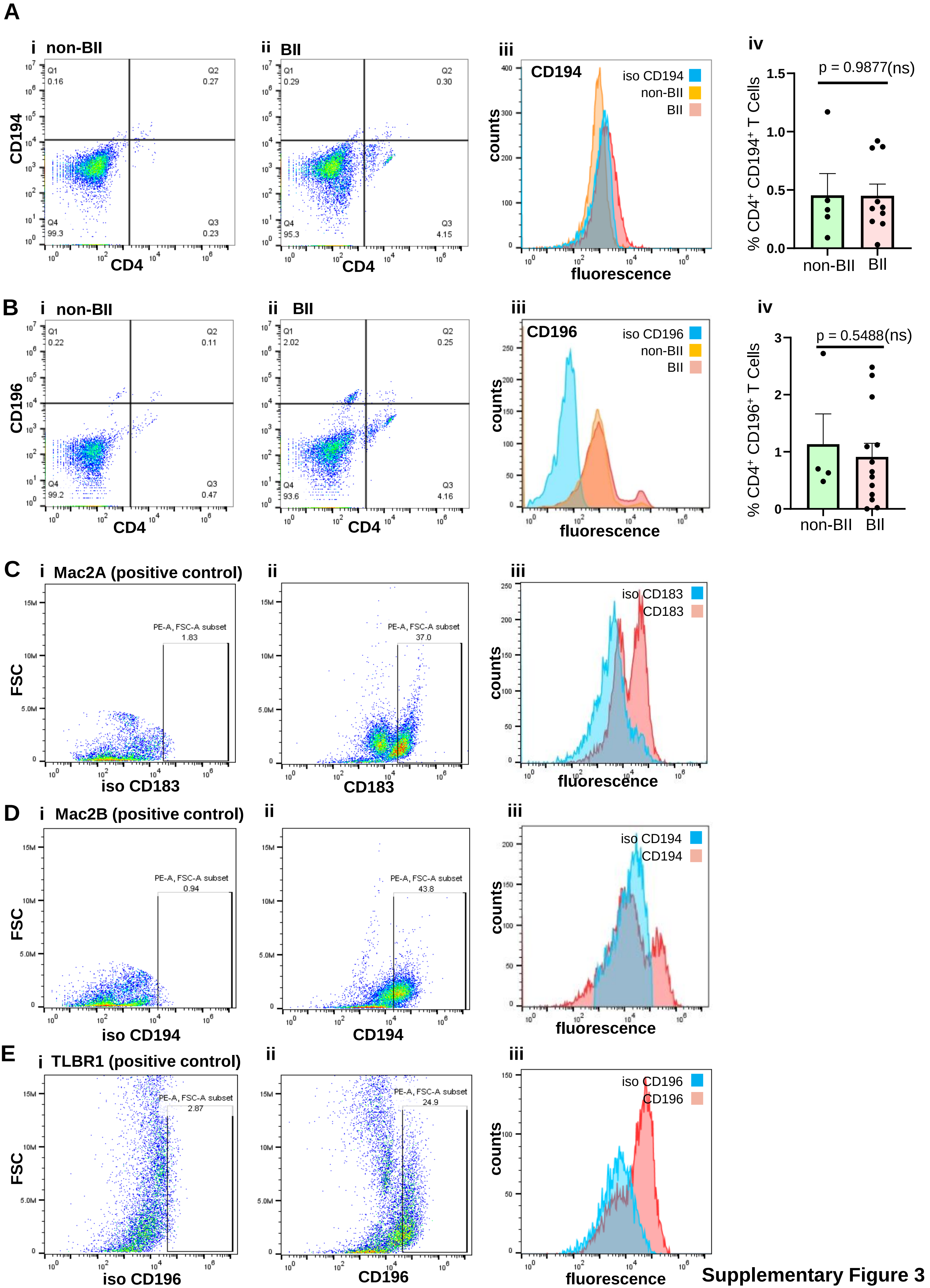

Supplementary Figure 3

**Supplementary Fig 3**

**(A)** Flow cytometry analyses of peripheral blood of subjects stained with anti-CD4 (FITC) and anti-CD194 (PE). Representative flow plots. (i) non-BII (ii) BII (iii) histogram with isotype control for CD194 (iv) % of $CD4^{+}$ $CD194^{+}$ T cells. Data presented as mean ± SEM, (n= 5-10).

**(B)** Flow cytometry analyses of peripheral blood of subjects stained with anti-CD4 (FITC) and anti-CD196 (PE). Representative flow plots. (i) non-BII (ii) BII subject (iii) histogram with isotype control for CD196 (iv) % of $CD4^{+}$ $CD196^{+}$ T cells. Data presented as mean ± SEM (n= 4-13).

**(C)** Flow cytometry of $CD4^{+}$ Mac 2A cell line (positive control) stained with isotype control to CD 183 or anti-CD183. Representative flow plots. (i) isotype control (ii) CD183 (iii) histogram with isotype control for CD183.

**(D)** Flow cytometry of $CD4^{+}$ Mac 2B cell line (positive control) stained with isotype control to CD 194 or anti-CD194. Representative flow plots. (i) isotype control (ii) CD194 (iii) histogram with isotype control for CD194.

**(E)** Flow cytometry of $CD4^{+}$ TLBR1 cell line (positive control) stained with isotype control CD 196 or anti-CD196. Representative flow plots. (i) isotype control (ii) CD196 (iii) histogram with isotype control for CD196.

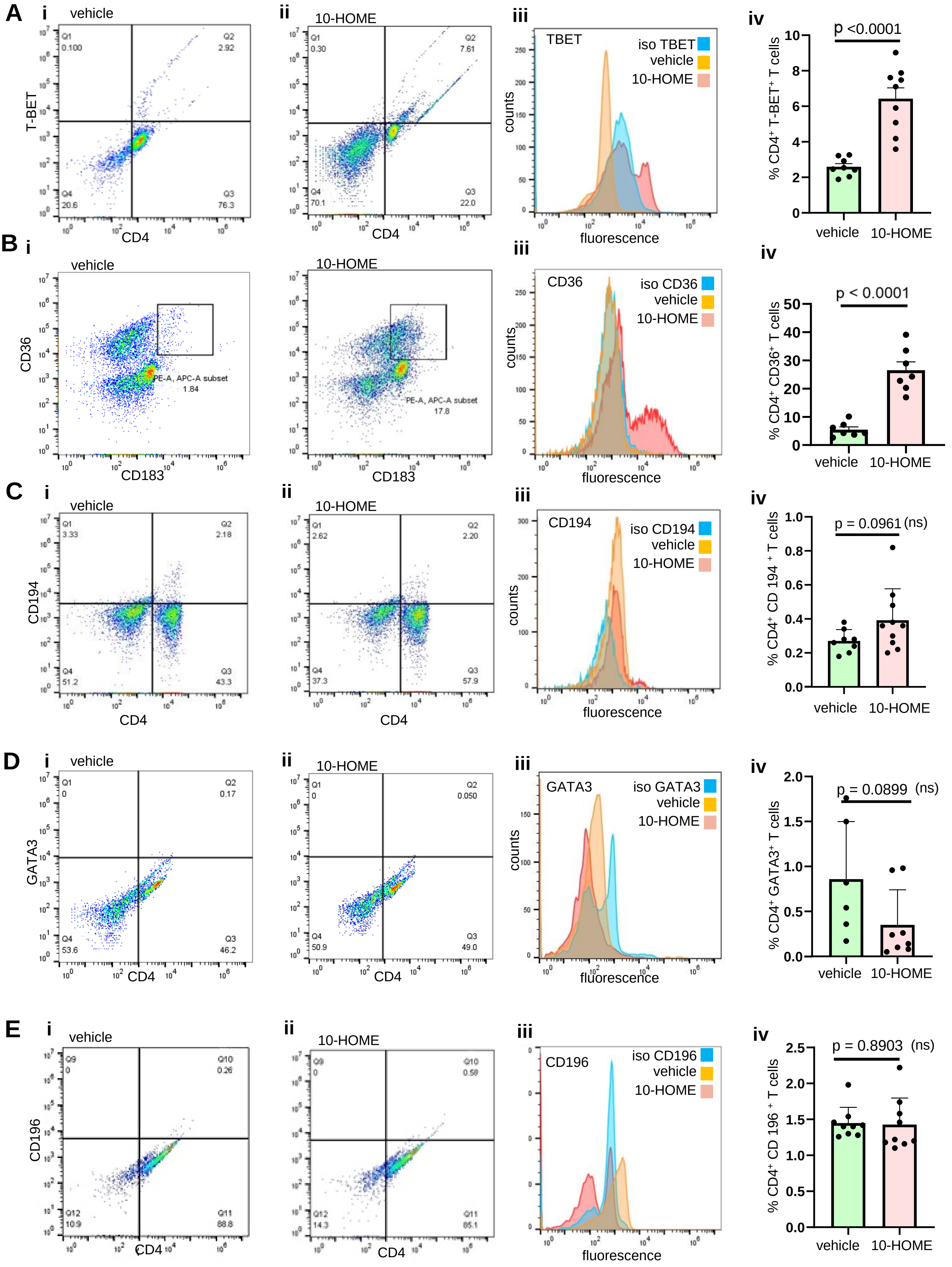

**Supplementary Figure 4**

**Supplementary Fig 4**

**(A)** Elevated TBET in the 10-HOME treated naïve $CD4^{+}$ T cells. Flow cytometry analyses of treated cells stained with anti-CD4 (FITC) and anti-TBET (PE). Representative flow plots. (i) vehicle treated (ii) 10-HOME treated (iii) histogram with isotype control for TBET (iv) % of $CD4^{+}$ $TBET^{+}$. Data presented as mean ± SD, (n=8-9).

**(B)** Elevated CD36 in the 10-HOME treated naïve $CD4^{+}$ $183^{+}$ T cells. Flow cytometry analyses of treated cells stained with anti-CD36 (APC) and anti-CD183 (PE). Representative flow plots. (i) vehicle treated (ii) 10-HOME treated (iii) histogram with isotype control for CD36 (iv) % of $CD4^{+}$ $TBET^{+}$. Data presented as mean ± SD, (n=7).

**(C)** Flow cytometry analyses of treated cells stained with anti-CD4 (FITC) and anti-CD194 (PE). Representative flow plots. (i) vehicle treated (ii) 10-HOME treated (iii) histogram with isotype control for CD194 (iv) % of $CD4^{+}$ $CD194^{+}$. Data presented as mean ± SD, (n=8-10).

**(D)** Flow cytometry analyses of treated cells stained with anti-CD4 (FITC) and anti-GATA3 (PE). Representative flow plots. (i) vehicle treated (ii) 10-HOME treated (iii) histogram with isotype control for GATA3 (iv) % of $CD4^{+}$ $GATA3^{+}$. Data presented as mean ± SD, (n=6-8).

**(E)** Flow cytometry analyses of treated cells stained with anti-CD4 (FITC) and anti-CD196 (PE). Representative flow plots. (i) vehicle treated (ii) 10-HOME treated (iii) histogram with isotype control for CD196 (iv) % of $CD4^{+}$ $CD196^{+}$. Data presented as mean ± SD, (n=9).

### Supplementary Table 1

**Breast Implant Illness Questionnaire**
**Approved through IRB # 2003674175**
**Title: Molecular Mechanisms Associated with Breast Implant Complications**

**Name:** ______________________________________________________________________
Date: ________________________

**Date of Birth:** ____/_______/_______ **Height:** _________ **Weight:**__________ Race/ **Ethnicity:** ____________

**Reason Implants Placed:** Augmentation Reconstruction **If reconstruction, were you diagnosed with breast cancer?** Y / N

**Date Implants Placed:** ____/_______/_______ **Original Surgeon:** __________________

**<u>Implant Information</u>**

**Manufacturer:** ________________________________ **Type:** Saline/ Silicone Gel Implant or Expander

**Texture:** Smooth Textured **Infection**: Clinically infected / Clean **Did Subject have Mastitis**: Yes / No

**Placement:** Above Muscle Below Muscle

**Incision Placement:** Transaxillary (underarm) Inframammary fold (breast crease) Periareolar (nipple) Other: _____________

**Implant Removed as:** Routine procedure / Due to self-reported complication of breast implant illness

**Is subject diabetic?** Yes / No **Subject received antibiotic irrigation:** Yes / No

**Have you had any other breast implant surgeries not listed above (e.g. breast lift, revision augmentation, biopsy):**
____________________________________________________________________________
_______________________________________________________

**When did your symptoms begin?**
____________________________________________________________________________
___________________________

**What were your initial symptoms?**______________________________________________

______________________________

**When/how did you become aware of Breast Implant Illness (BII)?**

________________________________________________________________

**<u>For office use only/Physician Notes:</u>**

________________________________________________________________

____________________

________________________________________________________________

______________________________________________

________________________________________________________________

______________________________________________

| | | | | | |
|---|---|---|---|---|---|
| **Local Chest Area** | | | | | |
| Do you experience pain or a burning sensation around the implant and/or the upper or outer chest? | Not at all | A little bit | Some what | Quite a bit | Very much |
| Do you experience pain and swelling in the armpit areas? | Not at all | A little bit | Some what | Quite a bit | Very much |
| Do you experience discomfort from tissue tightness, implant weight or pressure? | Not at all | A little bit | Some what | Quite a bit | Very much |
| **General** | | | | | |
| I feel fatigued. | Not at all | A little bit | Some what | Quite a bit | Very Much |
| I have brain fog, such as difficulty concentrating or memory loss. | Not at all | A little bit | Some what | Quite a bit | Very much |
| I have unexplained weight gain or loss *(circle which)*? How much? | Not at all | A little bit | Some what | Quite a bit | Very much |
| I have difficulty losing or gaining weight? | Not at all | A little bit | Some what | Quite a bit | Very Much |
| I feel inflamed. | Not at all | A little bit | Some what | Quite a bit | Very Much |
| I suffer from poor sleep. | Not at all | A little bit | Some what | Quite a bit | Very Much |
| I have foul body odor. | Not at all | A little bit | Some what | Quite a bit | Very Much |
| I feel much older than my true age. | Not at all | A little bit | Some what | Quite a bit | Very Much |
| **Immune System** | | | | | |
| I have frequent sinus infection. | Never | Rarely | Somet imes | Often | Always |
| I have frequent urinary tract infections. | Never | Rarely | Somet imes | Often | Always |
| I have frequent yeast infections. | Never | Rarely | Somet imes | Often | Always |
| I have frequent viral infections. | Never | Rarely | Somet imes | Often | Always |
| I have swollen or tender lymph nodes (armpit, neck). | Never | Rarely | Somet imes | Often | Always |
| I have unexplained frequent fevers. | Never | Rarely | Somet imes | Often | Always |
| I have chills. | Never | Rarely | Somet imes | Often | Always |
| I have night sweats. | Never | Rarely | Somet imes | Often | Always |
| **Psychological** | | | | | |
| I feel depressed. | Never | Rarely | Somet imes | Often | Always |

| | | | | | |
|---|---|---|---|---|---|
| I feel anxious. | Never | Rarely | Somet imes | Often | Always |
| I feel hopeless. | Never | Rarely | Somet imes | Often | Always |
| I have panic attacks. | Never | Rarely | Somet imes | Often | Always |
| **Musculoskeletal** | | | | | |
| I have joint pain and /or swelling?<br><br>If true, circle all that apply: Neck, shoulders, elbows, hands, back, hips, knees, feet | Never | Rarely | Somet imes | Often | Always |
| I have muscle pain and weakness. | Never | Rarely | Somet imes | Often | Always |
| I have muscle twitching. | Never | Rarely | Somet imes | Often | Always |
| I have slow muscle recovery after exercise. | Never | Rarely | Somet imes | Often | Always |
| **Pain Intensity** | | | | | |
| How would you rate your pain on average? (0 = no pain, 10 = worst pain imaginable)<br><br>1 2 3 4 5 6 7 8 9 10 | | | | | |
| **Skin** | | | | | |
| I have dry skin and hair. | Not at all | A little bit | Some what | Quite a bit | Very much |
| I have hair loss. | Not at all | A little bit | Some what | Quite a bit | Very much |
| I have skin rashes. Where? | Not at all | A little bit | Some what | Quite a bit | Very much |
| I have acne or acne-like eruptions. | Not at all | A little bit | Some what | Quite a bit | Very much |
| **Eyes** | | | | | |
| I have dry eye. | Not at all | A little bit | Some what | Quite a bit | Very much |
| I have vision changes or visual distortions. | Not at all | A little bit | Some what | Quite a bit | Very much |
| I have puffy eyes. | Not at all | A little bit | Some what | Quite a bit | Very much |
| I am sensitive to light. | Not at all | A little bit | Some what | Quite a bit | Very much |

| Respiratory | | | | | |
|---|---|---|---|---|---|
| I have a cough. | Not at all | A little bit | Some what | Quite a bit | Very much |
| I have chest congestion. | Not at all | A little bit | Some what | Quite a bit | Very much |
| I have shortness of breath. | Not at all | A little bit | Some what | Quite a bit | Very much |
| I have nasal discharge. | Not at all | A little bit | Some what | Quite a bit | Very much |
| **Heart** | | | | | |
| I feel palpitations. | Never | Rarely | Somet imes | Often | Always |
| I have chest pain. | Never | Rarely | Somet imes | Often | Always |
| **Gastrointestinal** | | | | | |
| I have food intolerance or allergies. | Never | Rarely | Somet imes | Often | Always |
| I have constipation. | Never | Rarely | Somet imes | Often | Always |
| I have diarrhea. | Never | Rarely | Somet imes | Often | Always |
| I have bloating. | Never | Rarely | Somet imes | Often | Always |
| I have abdominal pain. | Never | Rarely | Somet imes | Often | Always |
| I have reflux or gastritis. | Never | Rarely | Somet imes | Often | Always |
| I have dry mouth. | Never | Rarely | Somet imes | Often | Always |
| I have difficulty swallowing, a choking feeling, or a lump in my throat. | Never | Rarely | Somet imes | Often | Always |
| **Endocrine/Hormonal** | | | | | |
| I have temperature intolerance. | Never | Rarely | Somet imes | Often | Always |
| I have low libido. | Never | Rarely | Somet imes | Often | Always |
| I have heavy menstrual bleeding. | Never | Rarely | Somet imes | Often | Always |
| I have abnormal menstrual cycles. | Never | Rarely | Somet imes | Often | Always |
| I have symptoms of adrenal imbalance (slow healing, easy bruising). | Never | Rarely | Somet imes | Often | Always |

| Neurological | | | | | |
|---|---|---|---|---|---|
| I have metallic tastes. | Never | Rarely | Sometimes | Often | Always |
| I have dizziness. | Never | Rarely | Sometimes | Often | Always |
| I have ringing in my ears. | Never | Rarely | Sometimes | Often | Always |
| I experience numbness and tingling (e.g. arms, hands, fingers, legs, feet) | Never | Rarely | Sometimes | Often | Always |
| I have headaches (tension or typical). | Never | Rarely | Sometimes | Often | Always |
| I have migraines. | Never | Rarely | Sometimes | Often | Always |
| I have unusual facial or eye movements. | Never | Rarely | Sometimes | Often | Always |

**Do you feel your symptoms fall into one of these categories? If so, please check all that apply.**

☐Allergic ☐Autoimmune/Immune Disruption ☐Hormone Disruption ☐Neurologic ☐Mechanical (physical weight or tension from the implant) ☐Psychological (anxiety, regret, dissatisfaction)

**Do you have any of the following actual diagnoses (*check all that apply)***:

**Endocrine**: ☐Hypothyroid ☐Hyperthyroid ☐Hashimoto's ☐Grave's Disease ☐Infertility ☐Diabetes

**Pulmonary**: ☐Asthma

**Autoimmune**: ☐Rheumatoid arthritis ☐Lupus ☐Scleroderma ☐ Dermatomyositis ☐Sjogren's Syndrome ☐Multiple sclerosis ☐Nonspecific connective tissue disease ☐Sarcoidosis ☐Raynaud's ☐Interstitial cystitis

**Immune**: ☐Allergies ☐Immune deficiency ☐Reactivation of viruses such as EBV or Varicella (shingles)

**Musculoskeletal**: ☐Degenerative arthritis ☐Fibromyalgia

**Gastrointestinal**: ☐Irritable bowel syndrome ☐Ulcerative colitis ☐Crohn's colitis ☐SIBO

**Psychological**: ☐Depression ☐Anxiety

**Did you ever have mastitis? Yes No**

**Have you had a cholecystectomy (gallbladder removal)? Yes No *If yes, before or after implants?* Before After**

**Are you taking diabetic medications? Medication name(s):**

______________________________________________________________________

______________________________________________________________________
__________________________________________________

**Do you have a family history of autoimmune or connective tissue diseases? Yes No**

***If yes, please explain:***

______________________________________________________________________
________________________________

______________________________________________________________________
__________________________________________________

**Have you had any testing related to the symptoms you are experiencing? Yes No**

***If yes, please explain*:**

______________________________________________________________________
_________________________________________________

______________________________________________________________________
_________________________________________________

**Were any of the test results reported as abnormal? Yes No**

***If yes, please explain*:**

______________________________________________________________________
________________________________

______________________________________________________________________
__________________________________________________

**Name of physicians seen regarding symptoms:**

**Primary Care:**

______________________________________________________________________
_______________________________________

**OB/GYN:**

______________________________________________________________________
_________________________________________

**Functional Medicine:**

______________________________________________________________________________

____________________________________

**Rheumatologist:**

______________________________________________________________________________

_________________________________________

**Neurologist:**

______________________________________________________________________________

_____________________________________________

**Infectious Disease:**

______________________________________________________________________________

______________________________________

**Other:**

______________________________________________________________________________

__________________________________________________

**Any other history you would like to share: ______________________**

### Supplementary Table 2

| | Group | Size N | Median age | Mean duration of implant placed | Type of implant (smooth or textured) | Race/Ethnicity |
|---|---|---|---|---|---|---|
| 1 | BII | 46 | 44 | 11.5 | Smooth- 38<br>Textured- 8 | Caucasian -46 |
| 2 | non-BII | 14 | 46.5 | 6.1 | Smooth-10<br>Textured-4 | Caucasian -12<br>African American -2 |
| 3 | normal | 8 | 26.8 | not applicable | not applicable | Caucasian -6<br>African American-2 |

### Supplementary Table 3

| | Gene Name | Forward Primer | Reverse Primer |
|---|---|---|---|
| 1 | hTBET | CTC ACA AAC AAC AAG GGG GC | TCA CGG CAA TGA ACT GGG TT |
| 2 | hCD36 | ACT GAG GAC TGC AGT GTA GGA | AGT GGT TTC TAC AAG CTC TGG TT |
| 3 | hGAPDH | TGA CGC TGG GGC TGG CAT TG | GCT CTT GCT GGG GCT GGT GG |

Footnote h- human
